# An Evaluation of Empirical Approaches for Defining Cognitive Impairment in Amyotrophic Lateral Sclerosis

**DOI:** 10.1101/2020.12.23.20248410

**Authors:** Corey T. McMillan, Joanne Wuu, Katya Rascovsky, Stephanie Cosentino, Murray Grossman, Lauren Elman, Colin Quinn, Luis Rosario, Jessica H. Stark, Volkan Granit, Hannah Briemberg, Sneha Chenji, Annie Dionne, Angela Genge, Wendy Johnston, Lawrence Korngut, Christen Shoesmith, Lorne Zinman, the Canadian ALS Neuroimaging Consortium (CALSNIC), Sanjay Kalra, Michael Benatar

**Affiliations:** University of Pennsylvania Perelman School of Medicine, Department of Neurology, Philadelphia, PA, USA; University of Miami Miller School of Medicine, Department of Neurology, Miami, FL, USA; Columbia University, The Taub Institute for Research on Alzheimer’s Disease and the Aging Brain, New York, NY, USA; Division of Neurology, Department of Medicine, University of British Columbia, Vancouver, Canada; Department of Clinical Neurosciences, Hotchkiss Brain Institute, University of Calgary, Calgary, Canada; Department of Medicine, Université Laval, Québec, Canada; Montreal Neurological Institute and Hospital, McGill University, Montreal, Canada; London Health Sciences Centre, Western University, London, Canada; Sunnybrook Health Sciences Centre, University of Toronto, Toronto, Canada; Division of Neurology, Department of Medicine, University of Alberta, Edmonton, Canada; Neuroscience and Mental Health Institute, University of Alberta, Edmonton, Canada

## Abstract

**Importance:** Amyotrophic lateral sclerosis (ALS) is a multi-system disorder characterized primarily by motor neuron degeneration, but may be accompanied by cognitive dysfunction. Statistically appropriate criteria for establishing cognitive impairment (CI) in ALS are lacking.

**Objective:** Define thresholds for CI in ALS using quantile regression (QR) that accounts for age and education in a North American (NAmer) cohort.

**Design:** QR of cross-sectional data from a multi-center NAmer cohort of healthy adults was used to model the 5^th^ percentile of cognitive scores on the Edinburgh Cognitive and Behavioral ALS Screen (ECAS). The QR approach was compared to a traditional 2 standard deviation (SD) cut-off approach using the same NAmer cohort (2SD-NAmer) and to existing UK-based normative data derived using the 2SD approach (2SD-UK) to assess the impact of cohort selection and statistical model in identifying CI ALS patients.

**Participants:** 269 healthy adults from NAmer, recruited by the University of Pennsylvania (PENN; N=82), the University of Miami through the CRiALS study (CRiALS; N=40), and the Canadian ALS Neuroimaging Consortium (CALSNIC; N=147) were included to establish ECAS thresholds for defining CI. We then evaluated the frequency of CI in 182 ALS patients from PENN.

**Main Outcomes:** We defined two new sets of normative thresholds, based on NAmer heathy adult performance, for each ECAS domain score and the composite scores using QR and 2SD statistical approaches. We then applied the 2SD-NAmer and QR-NAmer, as well as the previously established and widely-used 2SD-UK, thresholds to evaluate the frequency of CI in ALS patients.

**Results:** QR-NAmer models revealed that increased age and reduced educational attainment negatively impact cognitive performance on the ECAS. Based on the QR-NAmer normative cutoffs, the prevalence of CI in the 182 PENN ALS patients was 15.9% for ECAS ALS-Specific and 15.4% for ECAS Total. These estimates are more conservative than estimates ranging from 15.4%-34.6% impaired based on 2SD approaches.

**Conclusions and Relevance:** This report establishes normative thresholds for using ECAS to identify whether ALS patients in the NAmer population have CI. The choice of statistical method and normative cohort has a substantial impact on defining CI in ALS.

**Key Points:** *Question:* How to define cognitive impairment (CI) in amyotrophic lateral sclerosis (ALS) using the Edinburgh Cognitive and Behavioral ALS Screen (ECAS)?

*Findings:* Age- and education-adjusted quantile regression (QR) yields thresholds for defining CI that differ meaningfully from those derived from parametric methods without age- and education-adjustment. Thresholds also differ between UK and North American cohorts. Applying our North American-based QR norms to an American ALS cohort at a single center identified CI based on ECAS performance in ∼16% patients, compared to 15.4%-34.6% patients using other approaches.

*Meaning:* The choice of statistical method and normative cohort has a substantial impact on defining CI in ALS.

---

Amyotrophic lateral sclerosis (ALS) is a multi-system disorder primarily characterized by progressive lower motor neuron (LMN) and upper motor neuron (UMN) degeneration^1^, but there is also increasing evidence for cognitive and behavioral impairment^2^. Indeed, current criteria for ALS frontotemporal spectrum disorder (ALS-FTSD) recommend two axes for the diagnosis of ALS including (I) defining the motor neuron syndrome and (II) defining neuropsychological deficits^**3**^. Cognitive impairment in ALS is an important clinical consideration given its association with poor prognosis including reduced survival and functional decline^**4**^.

The Edinburgh Cognitive and Behavioral Assessment Screen (ECAS) is an established brief neuropsychological assessment designed specifically for ALS that accommodates physical challenges (e.g., limited ability to speak or write) that potentially confound traditional neuropsychological testing^**5**^. Administration of the ECAS takes approximately 25 minutes to complete and uses a 136-point scale to evaluate several domains of cognition including language, executive functioning, and verbal fluency that are commonly believed to be most often affected in ALS, collectively contributing to an “ALS-Specific” composite score; as well as visuospatial and memory domains that are thought to be less frequently affected in ALS and contribute to an “ALS-Nonspecific” score. The ECAS has been validated against more extensive neuropsychological tests^**6**,**7**^ and multimodal studies, suggesting that ECAS performance relates to biologically plausible regional neuroimaging^8^ and neuropathological burden^9^ in ALS patients. While the ECAS has been translated into several languages^10-13^, normative data to empirically define cognitive impairments in North American ALS patients are lacking, and only rare prior studies have considered age and education when defining thresholds of cognitive impairment^14,15^. Moreover, prior approaches to defining impairment cutoffs on the ECAS have typically been based on relatively small series of healthy control data and have been based on statistical models that may violate important assumptions about the psychometric properties of ECAS scores.

In this report we present and compare 3 sets of normative thresholds for defining cognitive impairment, calculated based on different reference populations (N=40 single-center United Kingdom controls vs. N=269 multi-center North American controls) and statistical methods: the most common approach of “2 standard deviations below the mean” (2SD) vs. quantile regression with covariate adjustment (QR), as summarized in Table 1. Moreover, we illustrate how the choice of normative cutoffs impacts the classification of impairment (or not) in a cohort of 182 ALS patients.

**Table 1.** Study populations from which normative data were collected, statistical methods, and threshold strategies used to define cognitive impairment on the ECAS

|  | **2SD-UK** | **2SD-NAmer** | **QR-NAmer** |
| --- | --- | --- | --- |
| **Study population** | N=40<br>UK Controls | N=269<br>North American Controls |  |
| **Statistical method** | Mean ± 2 SD | Mean ± 2 SD | Quantile Regression |
| **Threshold strategy** | 2 SDs below the mean* | 2 SDs below the mean | 5<sup>th</sup> percentile, adjusting for age and education |

Note. SD = Standard deviation; \* Cutoff values previously published<sup>5</sup> ; NAmer=North American.

## Methods

### Control Participants and Sources of Normative Data

Data to establish normative ECAS performance were pooled together from three North American sources: the research cohort at the Penn Frontotemporal Degeneration Center and Comprehensive ALS Center at the University of Pennsylvania (PENN); the multi-center study conducted by the Clinical Research in ALS Biomarker study at the University of Miami (CRiALS); and the multi-center study conducted by the Canadian ALS Neuroimaging Consortium (CALSNIC) ^16^. This resulted in a total of 269 healthy adults; each source of data is described in detail below and demographic characteristics are summarized in Table 1.

#### PENN

82 community-dwelling healthy adults who self-report no history of neurological or significant psychiatric condition and were screened to have a Mini Mental Status Examination score of 27-30. The PENN research protocol was approved by an Institutional Review Board convened at the University of Pennsylvania and all participants provided written consent following an approved informed consent procedure.

#### CRiALS

40 healthy adults recruited from across the U.S. with no history of neurological disease, no significant psychiatric history, and either have no family history of ALS or FTD or do not carry the genetic mutation(s) known to cause ALS/FTD in their family. The CRiALS research protocol was approved by an Institutional Review Board convened at the University of Miami and all participants provided written consent following an approved informed consent procedure.

#### CALSNIC

147 healthy adults recruited across Canada and the U.S. and with no history of neurological disease, head trauma, or significant psychiatric history. The CALSNIC research protocol was approved by a Health Research Ethics Board convened at each of the participating universities including University of Alberta, University of British Columbia, Université Laval, University of Calgary, the Montreal Neurological Institute and Hospital at McGill University, Western University, and University of Toronto. All participants at each of these institutions provided written consent following an approved informed consent procedure.

Education was recorded differently across studies, and for simplicity in interpretation, we categorically defined education for statistical models as individuals who completed a four-year college degree or higher level of education (≥ College) or did not (< College).

### ALS Participants

To illustrate the impact of different ECAS normative cutoffs on identifying cognitive dysfunction in ALS, we evaluated 182 ALS patients at PENN who were diagnosed by a board-certified neurologist according to published consensus criteria^1^. Demographic and clinical characteristics are described in Online Supplementary Table 1. In addition to a clinical diagnosis of ALS, further inclusion criteria included completion of the ECAS along with three clinical assessments comprised of (1) a seated forced vital capacity (FVC) assessment of respiratory function; (2) the Revised ALS Functional Rating Scale (ALSFRS-R); and (3) Penn Upper Motor Neuron Score (PUMNS). All patients participated in an informed consent procedure approved by an Institutional Review Board convened at the University of Pennsylvania.

### ECAS

All healthy adult control participants and ALS patients completed the North American ECAS (Version 1) using published guidelines and instructions (https://ecas.psy.ed.ac.uk/ecas-international/#American) as described in detail elsewhere^5^. All evaluators were trained in the administration and scoring of the ECAS, which assesses the participants’ oral response performance in each of the 5 domains (language, fluency, executive, memory, and visuospatial function).

### Statistical Approaches

All statistical analyses were performed using R software (Version 3.5.1). Visual assessment of histograms and Q-Q plots for each domain as well as the composite scores on the ECAS suggested that most scores are not normally distributed (see Online Supplementary Figure 1) and Kolmogorov-Smirnov tests relative to a normal distribution centered on the mean of each score confirmed lack of normality for each domain (all p<0.1e^-8^). Therefore, we compare the most common approaches for defining cutoffs based on 2 standard deviations (2SD) below the mean across both sources of normative data relative to a Quantile Regression (QR) approach that does not make Gaussian assumptions and further accommodates adjustments for age and education.

We additionally summarize descriptive results using both parametric (e.g., mean) and non-parametric (e.g., median) statistics to facilitate comparison to previous reports.

### Methods for Defining Thresholds of Cognitive Impairment

#### Parametric UK Norms (2SD-UK)

The most widely applied approach for defining cognitive impairment on the ECAS is based on a previously established < -2 standard deviation cutoff relative to the mean in each domain and composite subscore from a sample of 40 Edinburgh-based healthy adults^5^.

#### Parametric North American (2SD-NAmer)

For comparison purposes we also implemented a < -2 standard deviation cutoff relative to the mean in each domain and the composite subscores, but using our sample of 269 North American healthy adults.

#### Quantile Regression North American (QR-NAmer)

Quantile regression has one key benefit over traditional parametric approaches for defining normative cutoffs^14,15^. It makes no assumption about the data distribution and is, therefore, more appropriate for skewed data distributions that may have floor or ceiling effects. In addition, it can account for heteroscedasticity in which the variance of an outcome (e.g., ECAS score) is not uniform across a factor (e.g., age or education), whereas the 2SD method does not allow for covariate adjustments except through stratification^17^. QR-NAmer cutoffs were calculated using the R quantregGrowth package^18^ for the 5^th^ percentile of performance in each ECAS domain and composite subscores for the 269 healthy adults.

## Results

### Normative ECAS Performance in Healthy Adults

The study population of 269 healthy adults (60% with at least a college education) included 150 females (56%) and ranged in age from 24 to 83, with a mean ± SD of 55.5 ± 11.5 years (Table 2). Median [interquartile range] of ECAS Total, ALS Specific and ALS Non-Specific scores were 115 [110-120], 87 [81-90] and 29 [27-31], with maximum possible scores of 136, 100 and 36 respectively (Table 2). Age- and education-adjusted estimates of 5^th^ percentile thresholds for cognitive impairment are summarized for all ECAS scores, including each cognitive domain, in Table 3 (see also Figure 1 and Online Supplementary Figure 2). Importantly, the QR-NAmer approach demonstrates a significant decline in healthy adult performance associated with age on each composite score (Figure 1, Table 3), with higher educational attainment (completing college) increasing the threshold for cognitive impairment by 1-4 points. Moreover, for individual cognitive domains, the 5^th^ percentile of performance also showed a slope associated with age for memory, fluency, executive and language performance, but not the visuospatial domain in which performance approached ceiling across all ages (Online Supplementary Figure 2).

**Table 2.** Summary demographics and performance on the Edinburgh Cognitive & Behavioral Screen (ECAS) from three cohort studies of healthy adults (N=269) from North America.

| Demographics | Overall |  | CALSNIC |  | CRiALS |  | PENN |  |
| --- | --- | --- | --- | --- | --- | --- | --- | --- |
| Total N (% Female) | 269 (56%) |  | 147 (54%) |  | 40 (70%) |  | 82 (52%) |  |
| Age, Mean (SD) | 55.5 (11.5) |  | 56.7 (9.9) |  | 50.2 (12.8) |  | 56.0 (13.0) |  |
| ≥ College Education, % | 60% |  | 50% |  | 80% |  | 70% |  |
| ECAS Performance | Mean (SD) | Median [IQR] | Mean (SD) | Median [IQR] | Mean (SD) | Median [IQR] | Mean (SD) | Median [IQR] |
| Language | 26.5 (1.8) | 27 [26, 28] | 26.4 (2.0) | 27 [26, 28] | 26.6 (1.7) | 27 [26, 28] | 26.7 (1.5) | 27 [26, 28] |
| Fluency | 18.6 (4.0) | 20 [16, 20] | 18.0 (4.3) | 20 [16, 20] | 18.9 (3.0) | 20 [18, 20] | 19.5 (3.8) | 20 [18, 22] |
| Executive | 39.8 (4.5) | 41 [38, 43] | 39.3 (4.9) | 41 [38, 43] | 39.9 (4.4) | 41 [38, 43] | 40.7 (3.6) | 42 [38, 43] |
| Memory | 17.1 (2.9) | 17 [15, 19] | 16.8 (3.0) | 17 [15, 19] | 18.2 (2.4) | 19 [16.75, 20] | 17.1 (2.7) | 17 [15, 18.75] |
| Visuospatial | 11.8 (0.5) | 12 [12, 12] | 11.8 (0.4) | 12 [12, 12] | 11.8 (0.5) | 12 [12, 12] | 11.8 (0.6) | 12 [12, 12] |
| Total | 113.9 (9.3) | 115 [110, 120] | 112.4 (10.5) | 114 [108, 120] | 115.4 (7.6) | 117 [110, 121] | 115.9 (7.3) | 116 [112, 122] |
| ALS Specific | 84.9 (8.1) | 87 [81, 90] | 83.7 (8.9) | 86 [80, 89] | 85.4 (6.6) | 87 [81, 9] | 86.9 (6.7) | 89 [84, 91] |
| ALS Non-Specific | 29.0 (3.0) | 29 [27, 31] | 28.7 (3.1) | 29 [27, 31] | 30.0 (2.4) | 31 [28, 32] | 29.0 (2.8) | 29 [27, 30.75] |

**Table 3.** Summary of ECAS cutoffs including two standard deviation approach with published United Kingdom values (2SD-UK) ^5^ and in the North American cohort using a two standard deviation (2SD-NAmer) and Quantile Regression (QR-NAmer) statistical approaches.

| ECAS Score | 2SD-UK | 2SD-NAmer | Education | QR-NAmer (Age) |  |  |  |  |  |  |  |  |
| --- | --- | --- | --- | --- | --- | --- | --- | --- | --- | --- | --- | --- |
|  |  |  |  | 35 | 40 | 45 | 50 | 55 | 60 | 65 | 70 | 75 |
| Language | 26 | 23 | < College | 24 | 24 | 24 | 24 | 24 | 23 | 23 | 22 | 22 |
|  |  |  | ≥ College | 24 | 24 | 24 | 24 | 24 | 23 | 23 | 22 | 22 |
| Fluency | 14 | 11 | < College | 12 | 12 | 12 | 11 | 11 | 9 | 8 | 6 | 5 |
|  |  |  | ≥ College | 14 | 14 | 14 | 13 | 13 | 11 | 10 | 8 | 7 |
| Executive | 33 | 31 | < College | 33 | 32 | 32 | 31 | 31 | 30 | 30 | 29 | 29 |
|  |  |  | ≥ College | 32 | 31 | 31 | 30 | 30 | 29 | 29 | 29 | 28 |
| Memory | 13 | 11 | < College | 14 | 13 | 13 | 12 | 11 | 11 | 10 | 9 | 9 |
|  |  |  | ≥ College | 15 | 15 | 14 | 13 | 13 | 12 | 11 | 11 | 10 |
| Visuospatial | 10 | 11 | < College | 11 | 11 | 11 | 11 | 11 | 11 | 11 | 11 | 11 |
|  |  |  | ≥ College | 11 | 11 | 11 | 11 | 11 | 11 | 11 | 11 | 11 |
| Total | 105 | 95 | < College | 102 | 101 | 99 | 98 | 96 | 94 | 92 | 90 | 87 |
|  |  |  | ≥ College | 106 | 105 | 103 | 101 | 100 | 98 | 96 | 93 | 91 |
| ALS Specific | 77 | 69 | < College | 74 | 73 | 72 | 71 | 70 | 68 | 67 | 66 | 65 |
|  |  |  | ≥ College | 77 | 76 | 75 | 73 | 72 | 71 | 70 | 69 | 67 |
| ALS Non-Specific | 24 | 23 | < College | 26 | 25 | 25 | 24 | 23 | 23 | 22 | 22 | 21 |
|  |  |  | ≥ College | 27 | 26 | 25 | 25 | 24 | 24 | 23 | 23 | 22 |

**Figure 1.**
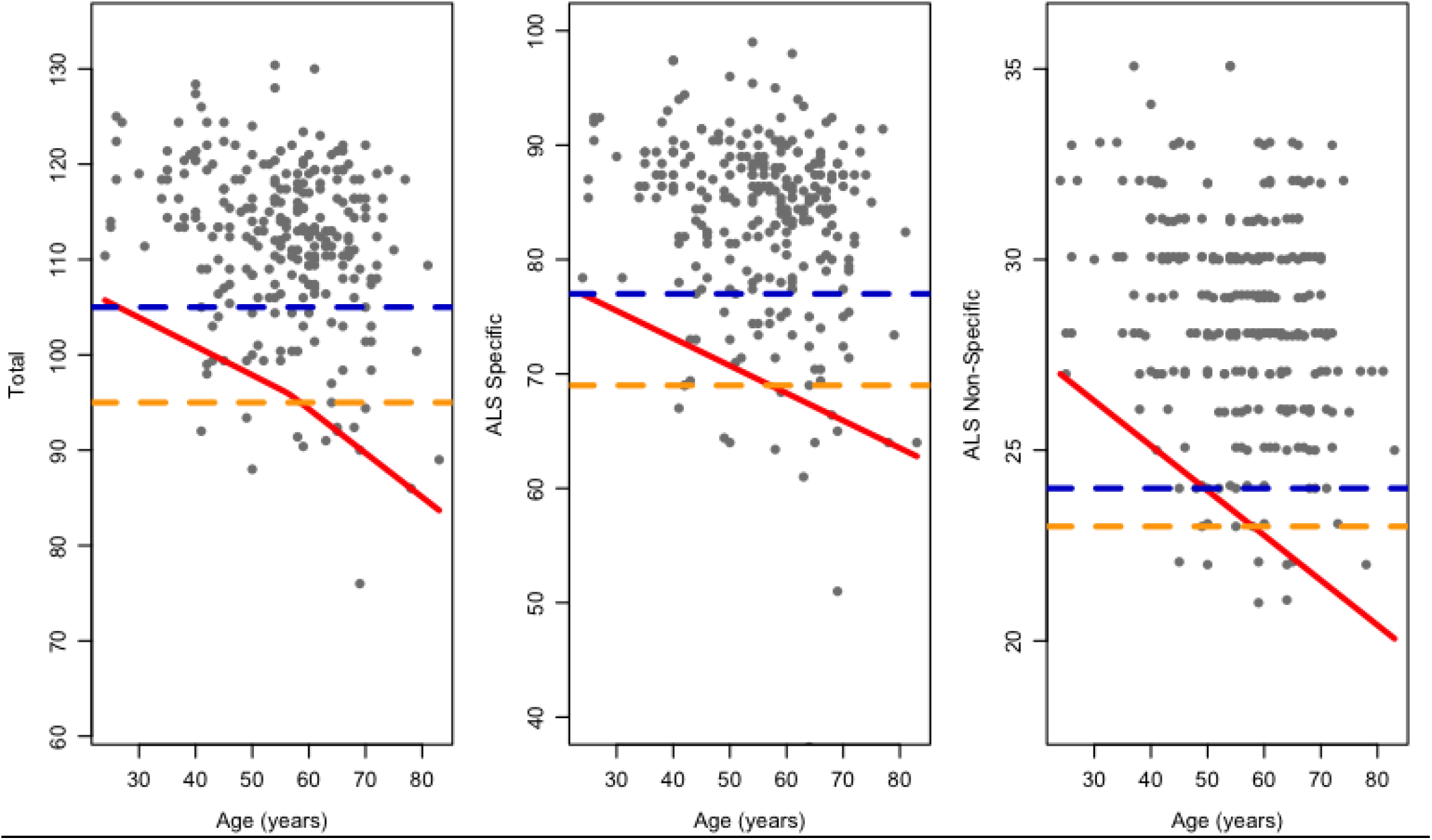
Quantile regression model for ECAS Total, ALS Specific, and ALS Non-Specific Scores across age, including 5^th^ Percentile cutoff (red) as well as for comparison the two standard deviation approach for North America (2SD-NAmer; orange) and United Kingdom (2SD-NAmer; blue).

### Prevalence of Cognitive Impairments in PENN ALS Cohort

The prevalence of cognitive impairment in ALS varies depending on the population used to generate normative data as well as the approach used to determine thresholds for defining cognitive impairment (Figure 2 and Online Supplementary Table 2). When comparing the statistical approaches (2SD and QR) developed across NAmer and UK cohorts, the 2SD-NAmer and QR-NAmer cutoffs suggest that ∼16% of PENN ALS patients exhibit cognitive impairment based on the ALS-Specific and ECAS Total composite scores, substantially fewer than that estimated using the 2SD-UK approach for ALS-Specific (32.4% impaired) or ECAS Total (34.6% impaired). The discordance between cutoff approaches is even more apparent in the Language domain with an estimated ∼63% impairment relative to ∼19% and ∼20% impaired in the PENN ALS cohort based on the 2SD-NAmer and QR-NAmer approaches, respectively. Fluency and Executive domains also suggest a lower frequency of cognitive impairment in the PENN ALS cohort using the 2SD-NAmer and QR-NAmer approach relative to the 2SD-UK approach.

**Figure 2.**
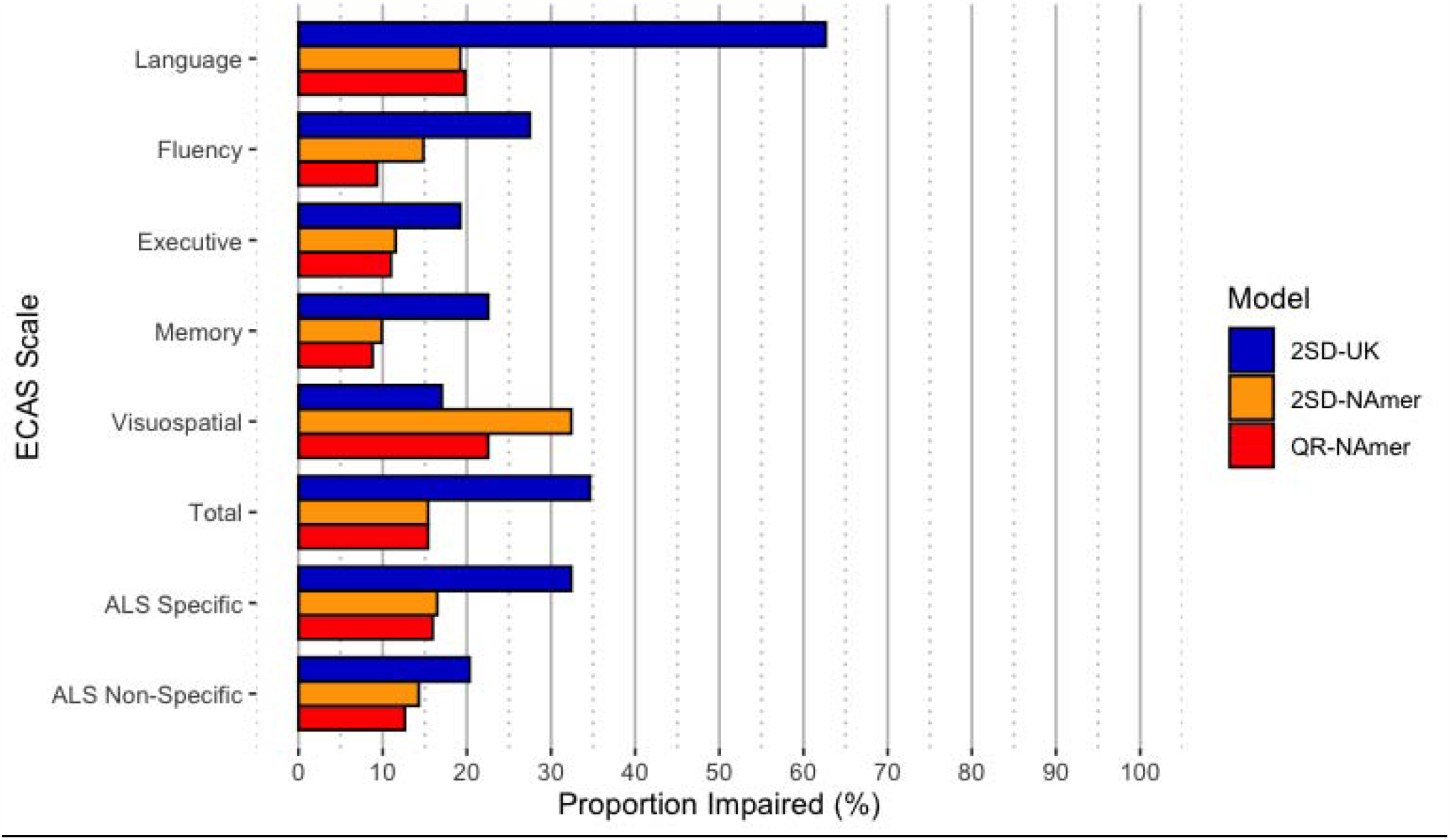
Prevalence of impairment on the ECAS in 182 ALS patients using three different sets of normative cutoffs. Note. QR-NAmer=Quantile regression North American; 2SD-NAmer=2 standard deviation cutoff North American; 2SD-UK= 2 standard deviation cutoff United Kingdom

In contrast to the NAmer vs. UK comparisons above, the differences between the QR and parametric statistical approaches yield relatively similar estimates when evaluating of the prevalence of cognitive impairment in the PENN ALS cohort using NAmer reference populations. These include the ALS Specific score - 15.9% (QR-NAmer) vs. 16.5% (2SD-NAmer) and the individual cognitive domains that contribute to this score: Language - 19.8% (QR-NAmer) vs. 19.2% (2SD-NAmer); Executive - 11.0% (QR-NAmer) vs. 11.5% (2SD-NAmer); and Fluency - 9.3% (QR-NAmer) vs. 14.8% (2SD-NAmer). The greatest divergence between QR-NAmer and 2SD-NAmer approaches was for Visuospatial impairment, 22.5% (QR-NAmer) vs. 32.4% (2SD-NAmer), that together with Memory (QR-NAmer=8.8% vs. 2SD-NAmer=9.9%) contributes to a discrepancy in rates of impairment on the ALS Non-Specific score - 12.6% (QR-NAmer) vs. 14.3% (2SD-NAmer).

## Discussion

Increasing interest in frontotemporal spectrum impairment in ALS and the overlap between ALS and FTD, have necessitated development of tools to quantify the nature and frequency of cognitive dysfunction in ALS. While the ECAS, a brief assessment tool, has emerged as the dominant instrument for evaluating cognition and behavior in ALS^19^, limited reference data (at least within North America) has hampered progress. In particular, efforts are underway to better understand genotype-phenotype relationships, including predictors of cognitive impairment^20^. Without appropriate normative data these efforts may be less successful due to potentially significant misclassification of cognitive impairment. Moreover, the recently revised Airlie House guidelines for the design and implementation of ALS clinical trials recommend that investigators stratify for cognitive impairments^21^; it is therefore essential to appropriately and accurately define whether individual patients are impaired or not.

Here we present normative data for the North American ECAS based on a moderately-sized, multi-center dataset and recommend (a) the use of quantile regression as appropriate for estimating normative thresholds (e.g. 5^th^ percentile) of data that are not normally distributed, and (b) adjusting for age and education, factors known to associate with cognitive function. Applying these findings to the ALS patient cohort at PENN, we have estimated the prevalence of cognitive impairment in ALS to be ∼16%. This is substantially less frequent than what we and others have previously estimated based on previously published norms, which suggested typically up to ∼50%^3,22^, and one estimate as high as 70%^23^, of ALS patients have cognitive impairment, though these prior estimates that are inflated in comparison to our observations were derived from neuropsychological tests that, unlike ECAS, do not control for motor confounds. Future work will be necessary to validate the QR-NAmer approach by comparing these suggested cutoffs relative to other more extensive neuropsychological assessments, but at present this is challenging because many other validated neuropsychological assessments are typically confounded by motor features.

While there is increasing evidence for language impairment in ALS, the current findings suggest that estimates of the rate of language impairments vary widely depending on the statistical approach and control population employed to generate normative data, ranging from ∼19% for the 2SD-NAmer and QR-NAmer approaches to as high as ∼63% of PENN ALS patients using the 2SD-UK approach. These findings therefore emphasize that sociodemographic differences across cultures are a critical consideration when defining normal language performance. As the ECAS becomes increasingly used internationally, it may be valuable to account for potential cultural and regional confounds in subsequent versions of the test, such as including cross-linguistic naming of objects ^24^, differences in orthographic-phonemic mapping that may influence spelling across languages, and implementing norms across several languages^25^. While letter-guided verbal fluency constitutes a “domain” distinct from language in the ECAS, it is linguistically-mediated and thus susceptible to cross-linguistic differences, such as variable letter frequency^26^. It will, therefore, likely be important to re-calibrate the “verbal fluency indices” across ECAS versions. Nevertheless, it is valuable to note the overall relatively high rate of impairment on language skills probed by ECAS. This has been noted by other investigators^27^, but may be underappreciated. Lastly, while semantic deficits have been reported in ALS^28^, there is increasing evidence for grammatical and discourse deficits in ALS ^29-31^. Supplementing the current language domain with tests of grammar and discourse in subsequent ECAS versions may provide a more accurate picture of language impairment in ALS.

Our use of the QR-NAmer approach, in addition to accounting for non-normality, benefits by also accounting for cognitive performance across age. Importantly, ECAS Total score cutoffs range from 106 to 87 points depending on the age of the individual, highlighting the importance of considering age in the determination of impairment by using a dynamic rather than static cutoff strategy. Memory appears to be the domain most affected by age, as well as the ALS Non-Specific score, which suggests that these components are more sensitive to age-related or non-FTD contributors to cognitive decline. Indeed, memory decline in aging may reflect mild cognitive impairment or prodromal Alzheimer disease (AD)^32^, and AD co-pathology has been noted in autopsy-confirmed series of ALS patients sensitive to cognitive impairment^33^. In this context, it is important to consider that ALS cognitive impairment (ALSci), as defined using current clinical criteria^3^, may be associated with the ALS-FTSD spectrum as a prodromal phase of ALS-FTD or ALS-Dementia. Importantly, the QR-NAmer approach suggests that the highest frequency of impairment based on composite scores (e.g., ALS Specific, ALS Non-Specific, Total) was the ALS Specific composite, which comprises the more predominantly affected executive, fluency, and language domains.

Beyond adjusting for age, the level of educational attainment has a modest impact on normative cutoff values. In particular, attainment of a college degree appears to have the greatest impact on verbal fluency and executive domains that contribute to the ALS Specific score, and only a 1-point impact on memory domains, while language and visuospatial performance remain constant across education groups. There are at least two possible reasons for this observation. One possibility for a lack of contribution of education to language and visuospatial domains may simply be a function of these domains having the most limited statistical variance. Another possibility, mentioned above, is that the limited range of probed language areas may be relatively insensitive to education. Yet an alternative possibility is that verbal fluency, executive, and memory domains are indeed preferentially impacted by educational status. For example, models of cognitive reserve suggest that factors like education may help stave off the clinical manifestations of underlying pathologies in aging individuals^34^ and evidence suggests that reserve may selectively impact executive functions but not visuospatial functions in healthy aging^35^ and frontotemporal degeneration^36^.

There are several caveats to consider. First, behavior is an important component of ALS-FTSD and, while this report establishes the frequency of cognitive impairment in ALS, it does not establish normative data for defining behavioral impairment in healthy adults or ALS patients. Second, our estimates of normative cognitive performance are based on a cross-sectional evaluation, which represents a snapshot of a progressive disease in which cognitive impairment may only emerge as disease unfolds^37^. While initial efforts have parametrically defined longitudinal change on the ECAS for healthy controls^38^, it is critical for future longitudinal investigations to define age-related rate of cognitive decline in healthy controls and in a manner that accounts for sociodemographic and cultural differences, in order to determine how the tempo of disease progression in ALS differs from normal age-related cognitive decline. Third, a limitation of pooling normative healthy adult data across cohorts is that educational attainment was not captured in a harmonized manner and therefore we were limited to dichotomizing education as college completion vs. less than a college education. However, cognitive performance may be more sensitive to finer-grained educational differences (e.g., high school, graduate degree) and this is an important consideration for future investigations. Likewise, our control cohort had a greater frequency of college completion (∼65%) than the general population; although the current study is the largest ECAS normative study to date, future studies with increased sampling of the general population may better capture the influence of education on ECAS performance. Lastly, we defined thresholds of cognitive impairment using a commonly implemented 5^th^ percentile cutoff for the QR approach and 2SD cutoff for the other approaches, but it is possible that alternative less conservative thresholds may be more appropriate.

In summary, estimates of the frequency of cognitive impairment in ALS vary widely depending on the cohort and statistical approach used to define thresholds for defining cognitive impairment. While our estimates are based on application of norms to ALS patients in the PENN cohort, we suggest that the use of quantile regression models, as employed in our QR-Namer approach, provides a robust strategy to define cognitive impairments that is statistically appropriate and accounts for potential confounding factors such as age and education.

## Data Availability

Data may be provided to outside investigators pending review of reasonable requests.

## Acknowledgments

We extend thanks to research coordinators at the University of Pennsylvania including Laura Baehr, Laura Hennessy, and Hayley Donaldson for their efforts with participant recruitment and evaluation. CALSNIC additionally thanks the research team for participant recruitment and evaluations. Additionally, CRiALS extends thanks to the research team at the University of Miami for participant recruitment and evaluation (Anne-Laure Grignon, Danielle Dauphin, Danielle Sheldon, Sumaira Hussain, Anne Cooley, Jessica Stark, Jessica Medina, Ashley Manso).

Dr. McMillan reports grants from the National Institutes of Health (NIH; AG017586; AG066597).

Dr. Kalra reports grants that support CALSNIC including the Canadian Institutes of Health Research, ALS Society of Canada, Brain Canada Foundation, and Shelly Mrkonjic ALS Research Fund.

Dr. Benatar reports grants from Muscular Dystrophy Association (Grants #4365 and #172123), the ALS Association (Grant #2015), and the National Institutes of Health (NIH; NS105479); philanthropic support from the ALS Recovery Fund and the Kimmelman Estate during the conduct of the study;

---

**Online Supplementary Table 1.**
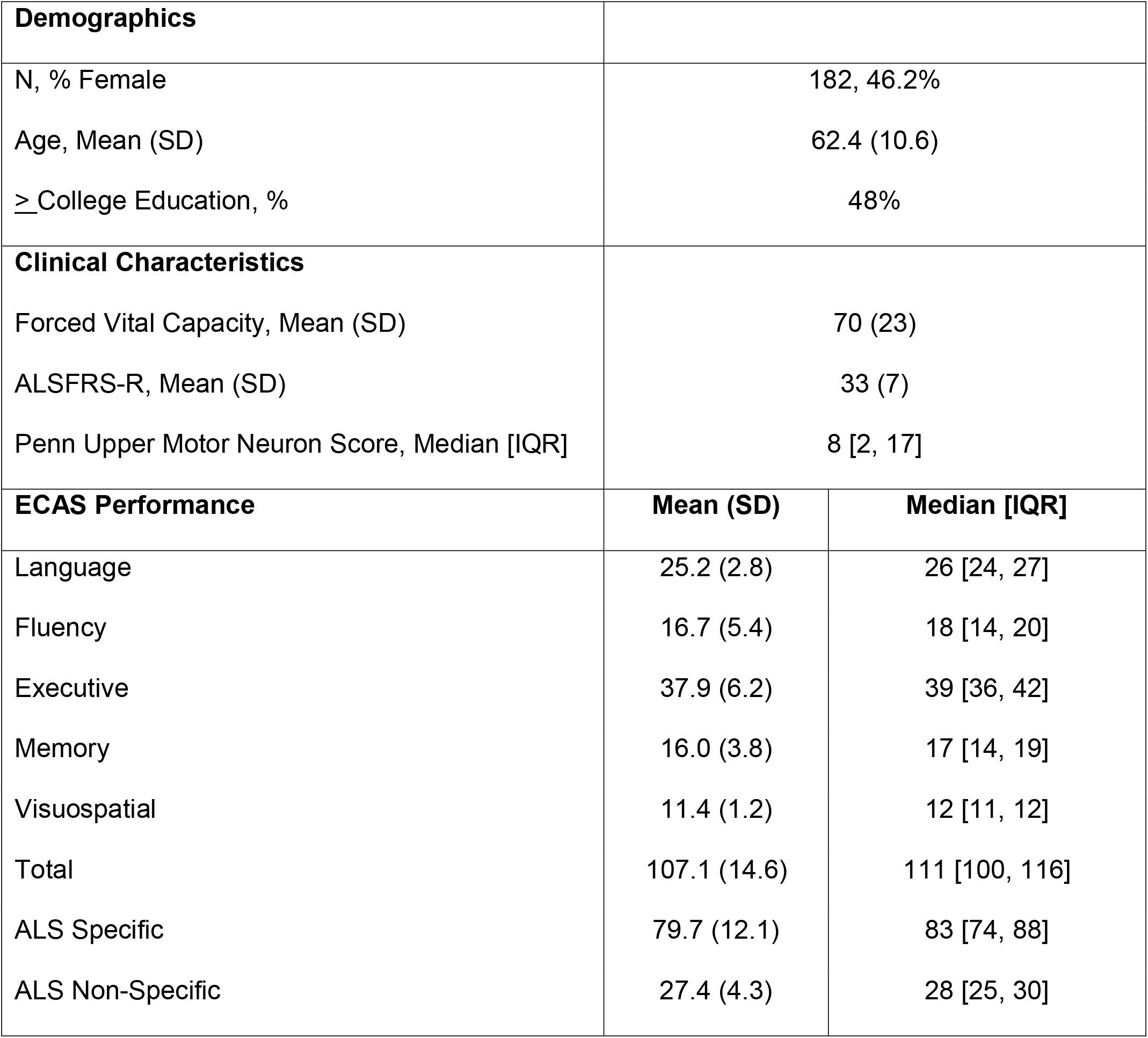
Summary demographics and ECAS performance for PENN ALS Patients (N=182)

**Online Supplementary Table 2.** Frequency (%) of impairment in 182 ALS based on cutoffs from two standard deviation cutoff approach from Abrahams et al. (2014, 2SD-UK), North American cohort (2SD-NAmer) cutoffs, and Quantile Regression North American (QR-NAmer) cutoffs.

| ECAS Score | 2SD-UK | 2SD-NAmer | QR-NAmer |
| --- | --- | --- | --- |
| Language | 114 (62.6%) | 35 (19.2%) | 36 (19.8%) |
| Fluency | 50 (27.5%) | 27 (14.8%) | 17 (9.3%) |
| Executive | 35 (19.2%) | 21 (11.5%) | 20 (11.0%) |
| Memory | 41 (22.5%) | 18 (9.9%) | 16 (8.8%) |
| Visuospatial | 31 (17.0%) | 59 (32.4%) | 41 (22.5%) |
| ECAS Total | 63 (34.6%) | 28 (15.4%) | 28 (15.4%) |
| ALS Specific | 59 (32.4%) | 30 (16.5%) | 29 (15.9%) |
| ALS Non-Specific | 37 (20.3%) | 26 (14.3%) | 23 (12.6%) |

**Online Supplementary Figure 1.**
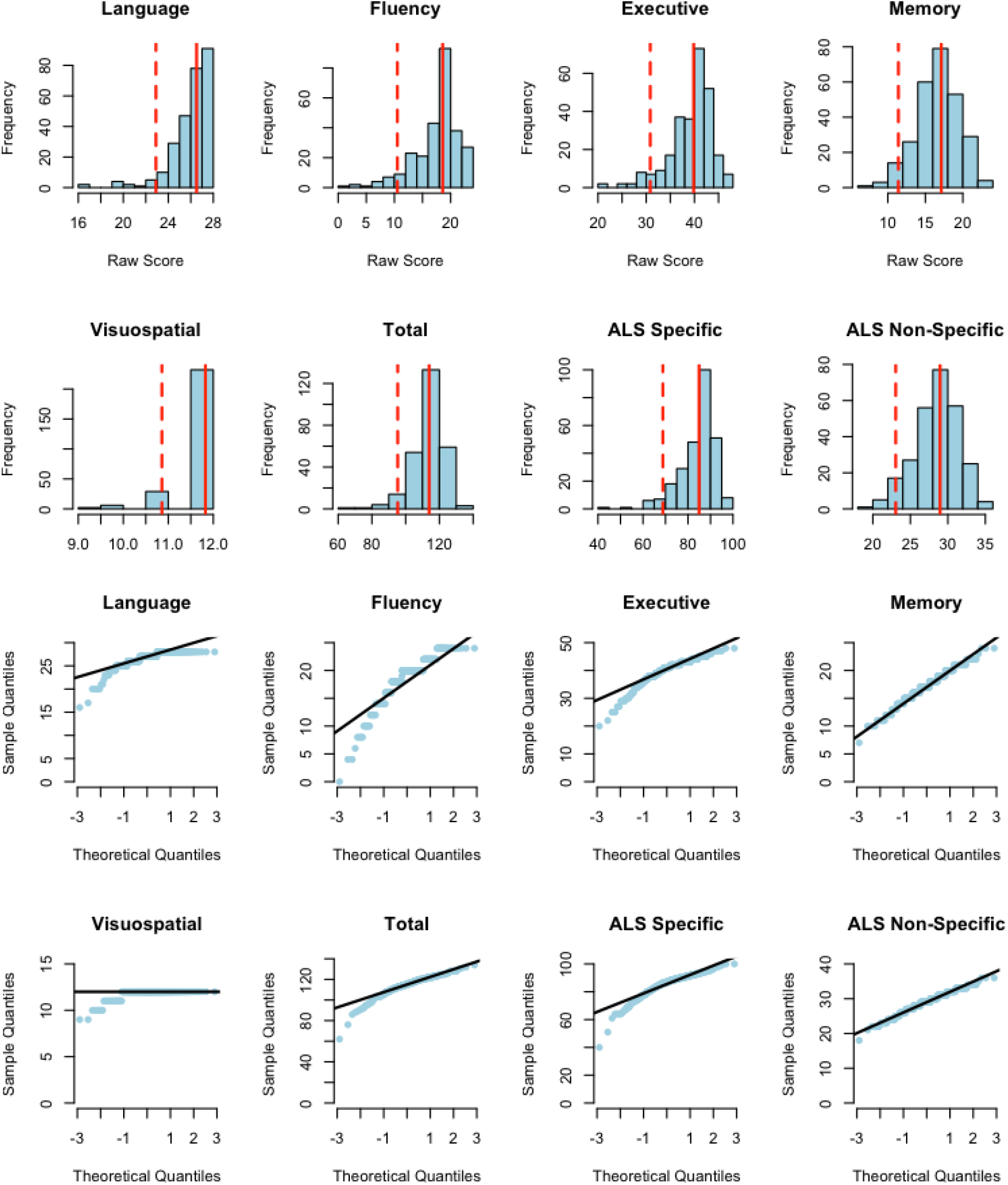
Histograms (top) and Quantile-Quantile (bottom) plots of the distribution of healthy adult performance on each ECAS domain and composite subscore. Note. In each histogram the solid red line represents mean performance and the dotted red line represents 2 standard deviations below the mean, selected to represent the threshold for impairment employed using the 2SD-NAmer approach.

**Online Supplementary Figure 2.**
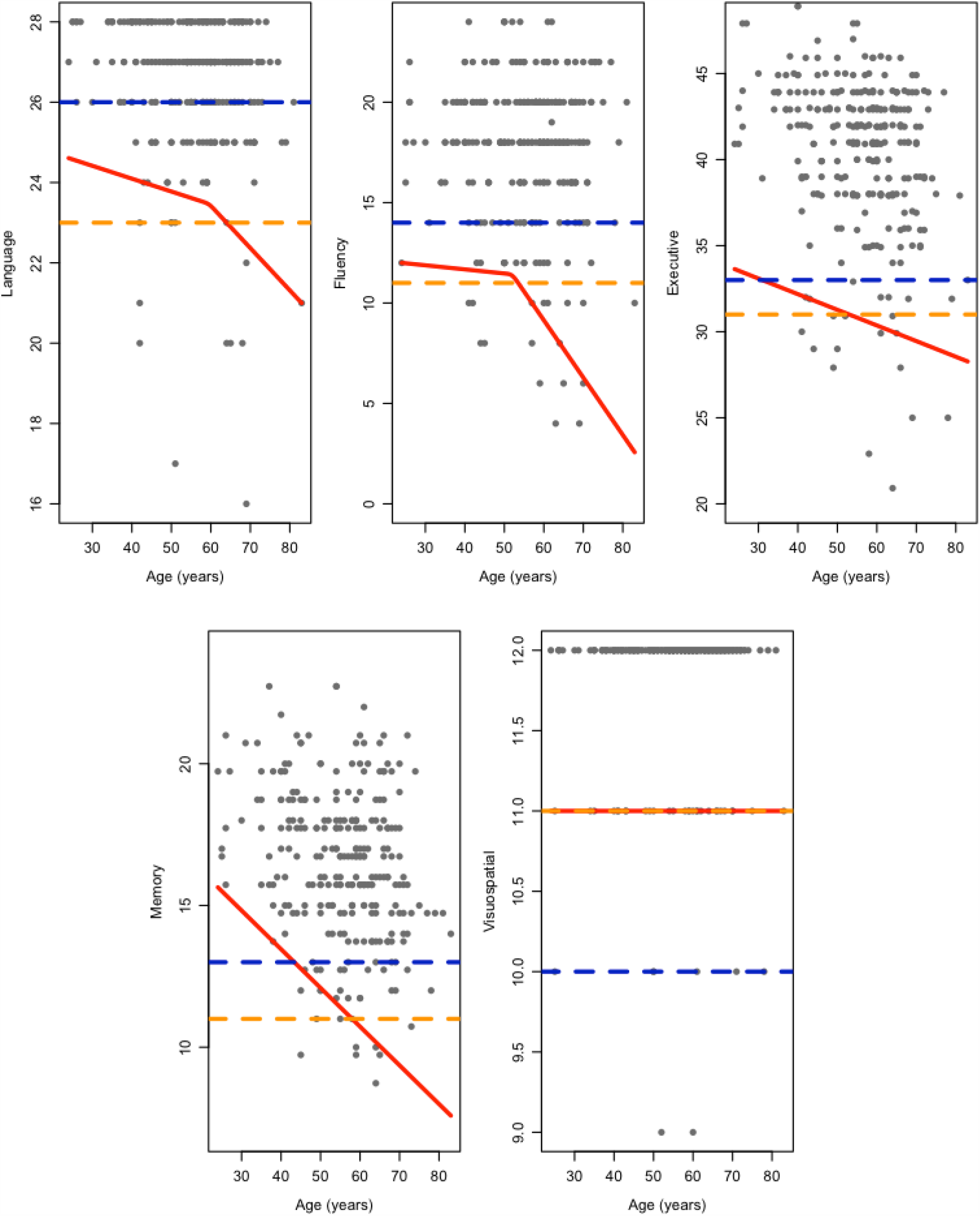
Quantile regression model for Language, Fluency, Executive, Memory, and Visuospatial domains, including 5^th^ Percentile cutoff (red) as well as for comparison the two standard deviation approach for North America (2SD-NAmer; orange) and United Kingdom (2SD-NAmer; blue).

